# Description of the Intervention for Virological Suppression in Youth with HIV (iVY): A telehealth behavioral intervention focused on mental health, substance use, and HIV care engagement among youth living with HIV

**DOI:** 10.64898/2026.07.13.26357907

**Authors:** Caravella McCuistian, Celeste Balaban, Hiromi Ortega Roque, Valerie A. Gruber, Mallory O. Johnson, Parya Saberi

**Author notes:** These authors contributed equally to this work (Caravella McCuistian, Celeste Balaban). Trial Registration: ClinicalTrials.gov NCT05877729 (https://clinicaltrials.gov/study/NCT05877729). Protocol and statistical analysis plan: https://pubmed.ncbi.nlm.nih.gov/37802624/. Data sharing: N/A. Funding: This work was supported by the National Institute of Mental Health of the National Institutes of Health under award number R01MH131415.

## Abstract

**Objective:** Youth with HIV experience persistent disparities across the HIV care continuum, including low rates of engagement in care and viral suppression. In a recent national survey, youth and young adults, defined by the CDC as ages 13–34, accounted for approximately 20– 40% of new HIV diagnoses in the United States. We describe the Intervention for Virological Suppression in Youth with HIV (iVY), a youth-friendly, tailored approach that integrates mental health and substance use support with HIV treatment engagement.

**Trial Design:** This paper describes the development of the intervention used in iVY, which is currently being evaluated in a randomized clinical trial (RCT) using an adaptive treatment strategy. HIV virological suppression is measured via dried blood spot at 16 weeks.

**Methods:** The intervention is delivered fully remotely across California and Florida. YWH aged 18–29 who are not durably virally suppressed are enrolled and randomized to the intervention or usual care. The RCT will enroll and randomize 200 participants to the intervention (*n* = 100) versus usual care (*n* = 100). iVY includes: (1) tailored brief, weekly video-counseling sessions focused on HIV treatment adherence and engagement, mental health, substance use, and related barriers; and (2) a mobile health application designed to support adherence, resource access, and peer connection. Participants who are not virally suppressed receive an additional 16 weeks of intensified intervention, while responders continue with app-based support.

**Results:** N/A

**Conclusion:** This paper provides a detailed description of a telehealth-based behavioral intervention tailored to the needs of youth with HIV. The intervention offers a scalable model for integrating behavioral health and HIV care to address barriers to treatment engagement in this priority population.

## Introduction

According to the 2022-2025 United States National HIV/AIDS Strategy, specific populations continue to experience disparate HIV incidence, despite a nationwide decline in prevalence [1,2]. Youth and young adults are identified as a priority population, accounting for over 7000 (or 19%) of new HIV diagnoses in 2022 [3]. Youth with HIV (YWH) also experience decreased treatment retention and viral suppression compared to other age groups [4,5], which is often compounded by mental health (MH) and substance use (SU) challenges.

Even though interventions to increase HIV treatment adherence for YWH exist [8–10], these interventions should be tailored to the unique needs of YWH [11], including MH and SU challenges, which can impact treatment for YWH across the care cascade [12–14]. Interventions that also consider YWH’s interest in technology-based delivery methods may be one solution to improving HIV treatment adherence [15].

The Intervention for Virologic Suppression in Youth (iVY) is a technology-based intervention that aims to address HIV treatment adherence, MH, and SU among YWH. The efficacy of iVY is currently being evaluated in a randomized clinical trial (RCT) with participants aged 18-29 years who are not durably virally suppressed. The trial aims to evaluate whether iVY improves HIV care engagement, virologic suppression, and mental health and substance use outcomes while monitoring the minimal risks associated with participation, including potential emotional discomfort related to discussing sensitive topics and privacy considerations inherent to telehealth delivery. This paper outlines the development of iVY and provides a detailed description of the intervention. Results of the ongoing trial’s efficacy are forthcoming.

## Methods

### Trial Design

The iVY study is an ongoing two-arm randomized clinical trial with a 1:1 allocation ratio comparing the iVY intervention with usual care. The trial incorporates an Adaptive Treatment Strategy (ATS), in which predefined decision rules individualize intervention intensity based on participants’ virologic response at 16 weeks. Participants who do not achieve virologic suppression receive an additional 16 weeks of intensified video counseling and app support, whereas participants who achieve virologic suppression continue with app-only support.

The sample size was based on power calculations for the primary outcome of virologic suppression. Assuming 20% attrition, 80% power, a two-sided α of 0.05, and a control-group virologic suppression rate of 44%, the study was powered to detect a clinically meaningful difference in virologic suppression between study arms. Detailed descriptions of the randomization procedures, allocation concealment, blinding, statistical analysis plan, and sample size determination are available in the published trial protocol [16].

### Intervention Overview

iVY is a 16-week intervention aimed at addressing MH and SU challenges for YWH to improve HIV treatment adherence and viral suppression. iVY combines individual video counseling with a mobile app (WYZ) to address barriers to treatment adherence. While iVY aims to address MH and SU, WYZ deals with medication management, community resources, and general HIV knowledge. iVY begins with 12 brief 20–30-minute one-on-one video counseling sessions delivered over a 16-week timeframe, allowing for missed sessions. These video counseling sessions were developed by the authors through formative research with youth living with HIV, input from the Youth Advisory Panel (YAP), interdisciplinary collaboration, and community involvement to improve HIV care engagement and treatment adherence and address MH and SU challenges among YWH. A pilot RCT of the video-counseling component of iVY was completed [17], with results showing high levels of video-counseling session completion, feasibility, acceptability, and satisfaction. This pilot RCT also revealed improved antiretroviral therapy (ART) adherence and HIV knowledge, decreased MH symptoms, and decreased MH and SU-related stigma [17]. Details on the development of the 12 video-counseling sessions offered in iVY are described elsewhere [18].

To augment the first 16 weeks of video-counseling sessions, iVY also provides access to the WYZ app. Designed using a human-centered design approach, WYZ comprises three separate sections (My Health, My Team, and My Community) that support the user in tracking their medication adherence and understanding their health, connecting with community resources, and receiving peer support from other YWH [19]. A pilot study of WYZ showed high feasibility, acceptability, and satisfaction [20].

In addition to combining features from the first 16-week intervention and WYZ, the iVY study includes a component called “focused sessions.” Delivered to individuals who remain virally unsuppressed at the end of the first 16 weeks of sessions, the focused sessions are an additional 12 sessions using behavioral techniques to augment skills introduced in the previous 12 sessions. Focused session topics and behavioral techniques were adapted from evidence-based treatment approaches (e.g., Dialectical Behavior Therapy, Cognitive Behavior Therapy, Motivational Interviewing). The intervention is described in accordance with the TIDieR checklist (Table 1), with further details provided below.

**Table 1.**
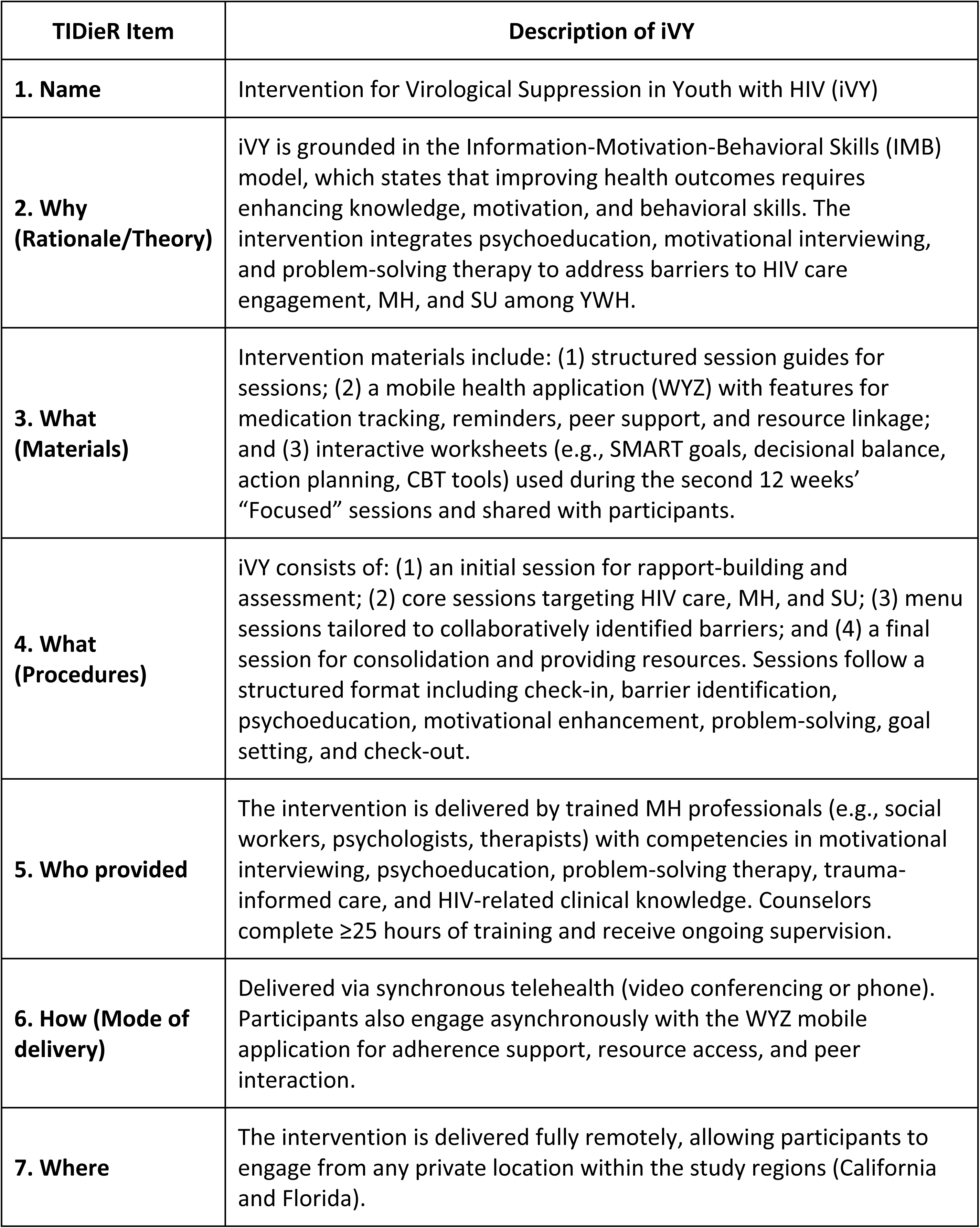
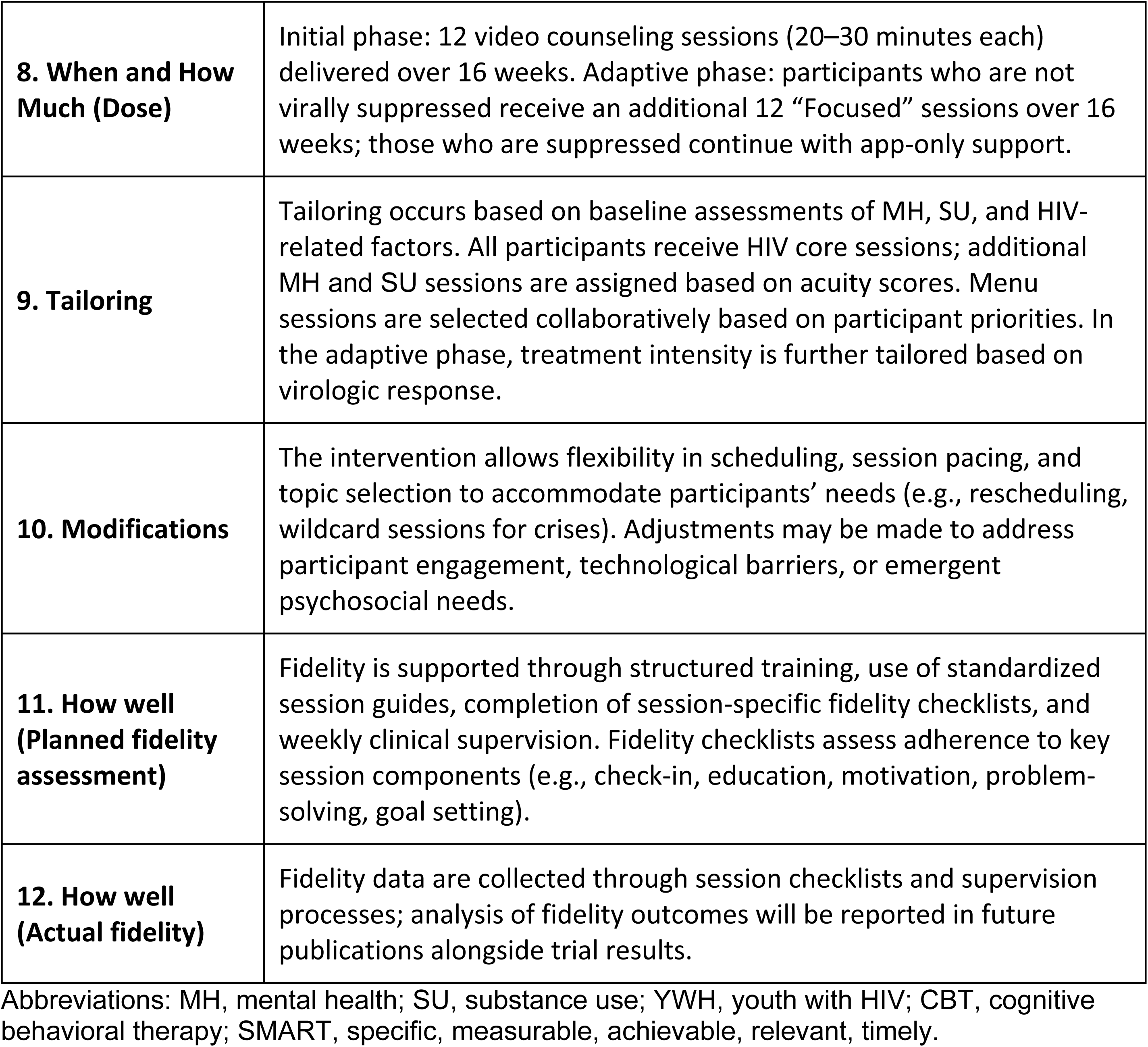
TIDieR Checklist for iVY.

### Participants

Study participants in iVY are primarily recruited through a partnership with AIDS Healthcare Foundation (AHF), a US non-profit organization providing HIV primary care services. Recruitment is also conducted via social media ads, dating apps, outreach to eligible former study participants, and distribution of flyers at events, festivals, and HIV primary care clinics.

Study inclusion criteria include English-speaking YWH ages 18-29 who have a documented detectable viral load (≥20 copies/mL) within the past year. All participants provided informed consent before participating in the study procedures. Written informed consent was obtained during a video-based enrollment visit, during which study staff reviewed the consent form verbally and provided participants with an opportunity to ask questions. The iVY randomized clinical trial is registered at ClinicalTrials.gov (NCT05877729; https://clinicaltrials.gov/study/NCT05877729). Participant recruitment occurred from 29/11/2023 to 31/12/2025. Follow-up assessments are conducted through 48 weeks after enrollment. The estimated study completion date is 04/24/2026.

### Theoretical Framework

iVY is informed by the information-motivation-behavior skills (IMB) model [21]. This model emphasizes the importance of providing information, enhancing motivation, and building behavioral skills as key approaches to encouraging behavior change. The IMB model suggests that providing education about HIV, increasing motivation for self-care, and teaching behavioral skills improve health, may result in improved engagement in HIV treatment [22].

iVY uses several techniques to address each factor of the IMB model. Psychoeducation, health education, or educating participants about their mental and physical health, is used to address the information component of the IMB model [23]. Motivational Interviewing, a counseling approach focused on eliciting and enhancing motivation towards behavior change, is used to address the motivation component [24]. Finally, problem-solving therapy assists individuals in addressing current stressors [25].

### Detailed Description of iVY Counselor Prerequisites and Training

iVY is delivered by a trained MH professional (e.g., clinical psychologist, social worker, therapist), referred to as the “counselor.” The counselor should be competent in psychoeducation, motivational interviewing, problem-solving, trauma-informed, and culturally sensitive therapeutic approaches. They are also trained in HIV-related clinical and psychosocial issues. Ideally, the counselor reflects the demographics of the participants in iVY (e.g., age, gender, race/ethnicity, or lived experiences).

Before delivering the intervention, counselors undergo a minimum of 25 hours of training alongside a current or former iVY counselor. The training is structured into five distinct phases: (1) general orientation, (2) introduction to session outlines, (3) session demonstrations and paired practice, (4) session review and feedback by supervisor, and (5) ongoing supervision and support. This rigorous training is designed to ensure fidelity to the intervention and to equip counselors with the skills necessary for effective implementation. Once the counselor begins working with participants, a fidelity checklist (see Table 2) is completed at the end of each session, and weekly supervision with licensed MH clinicians is provided. Additional training is encouraged. Examples of trainings completed by counselors include Motivational Interviewing for HIV Care Providers, Digital Mental Health and LGBTQIA+ Youth, Immigrant Mental Health, and Acceptance and Commitment Therapy (see <u>supplemental materials</u>). Potential harms were assessed systematically throughout the study through counselor monitoring during sessions, documentation using fidelity forms following each session, weekly clinical supervision, and implementation of predefined crisis and referral protocols when safety concerns were identified.

**Table 2.** Description of Focused Intervention Topics.

| **Focused Session Topic** | **Description** | **Motivation**<br>“This topic enhances motivation by...” | **Behavioral Skills**<br>“This topic builds behavioral skills by...” |
| --- | --- | --- | --- |
| **Core 1: Decisional Balance** | Evaluate the costs and benefits of health-related behavior change. | normalizing ambivalence and increasing participants’ insight into how current behaviors align with values and goals. | evaluating pros/cons of change and selecting aligned next steps. |
| **Core 2: SMART Goal** | Develop a clear and realistic health-related goal. | connecting goal to health benefits to strengthen confidence and readiness for change. | breaking larger barriers into measurable and actionable steps. |
| **Core 3: Action Plan** | Create a step-by-step plan to support a health-related goal. | increasing commitment through identification of supports and obstacles. | developing task-based plans and identifying practical strategies. |
| **Core 4: Monitoring and Evaluating Outcomes** | Review progress and modify goals as needed. | reinforcing confidence through reflection and refinement. | tracking outcomes, identifying barriers, and revising plans. |
| **Core 5: 7 Steps of Problem Solving** | Apply a structured problem-solving process to a health-related barrier. | reducing feelings of being stuck and increasing self-efficacy. | defining problems, generating solutions, and implementing plans. |
| **Menu A: Pleasurable Activities** | Identify and schedule pleasurable and mastery-based activities. | by increasing positive expectancy through meaningful and achievable activities. | scheduling activities and monitoring effects on mood and functioning. |
| **Menu B: Stress and Coping** | Identify stressors and apply coping strategies. | increasing awareness of and strengthening self-efficacy for managing stress-related barriers to health. | matching stressors with emotion-focused and problem-focused coping skills. |
| **Menu C: Effective Communication** | Practice assertive communication strategies related to health goals. | increasing confidence in communicating needs effectively. | applying assertive communication techniques in real-life situations. |
| **Menu D: Boundaries with Self and Others** | Practice values-based boundaries and safer coping responses. | strengthening confidence in setting healthy boundaries. | identifying triggers and practicing alternative coping responses. |
| **Menu E: Practicing Acceptance** | Use acceptance-based coping for unchangeable stressors. | increasing willingness to tolerate distress and uncertainty. | practicing acceptance-based self-talk and coping strategies. |
| **Menu F: CBT Thought-Emotion-Behavior Connection** | Identify interactions between thoughts, emotions, and behaviors. | Reducing feelings of overwhelm through increased self-awareness. | Mapping triggering situations and generating alternative responses. |
| **Menu G: Identifying Values** | Connect personal values with health-related goals. | connecting daily actions to core values to increase purpose, consistency, and alignment. | developing values-based goals and routines. |
| **Menu H: Recognizing and Responding to Triggers** | Identify triggers and develop values-based responses. | increasing readiness to respond differently to triggers. | identifying triggers and planning alternative responses. |
| **Menu I: Motivation Enhancement** | Explore readiness, values, and barriers to engagement. | enhancing change talk and motivation for engagement. | using motivation rulers and collaborative goal exploration. |

### Enrollment Visit

iVY begins with an enrollment visit conducted by a research coordinator over a HIPAA-compliant video-conferencing tool or by telephone. Informed consent, baseline surveys, training on using a home test kit to assess viral load at the end of the first 16 weeks of the intervention, and WYZ app orientation occur during this visit.

### Assessment, Acuity, and Intervention Tailoring

iVY uses responses to baseline surveys to assess symptom acuity and tailors the intervention for each participant (Fig 1). During enrollment and at 16, 32, and 48 weeks, participants complete assessments of MH, SU, and HIV knowledge and treatment adherence using validated measures, including the Patient Health Questionnaire-8 (PHQ-8) [26,27], Generalized Anxiety Disorder Scale-7 (GAD-7) [28], Post-traumatic Stress Disorder Checklist (PCL-5) [29], Alcohol Use Disorders Identification Test (AUDIT) [30], Drug Abuse Screening Test (DAST) (5), Alcohol, Smoking, and Substance Involvement Screening Test (ASSIST) [31], and the HIV Treatment Knowledge Scale [32].

**Fig 1.**
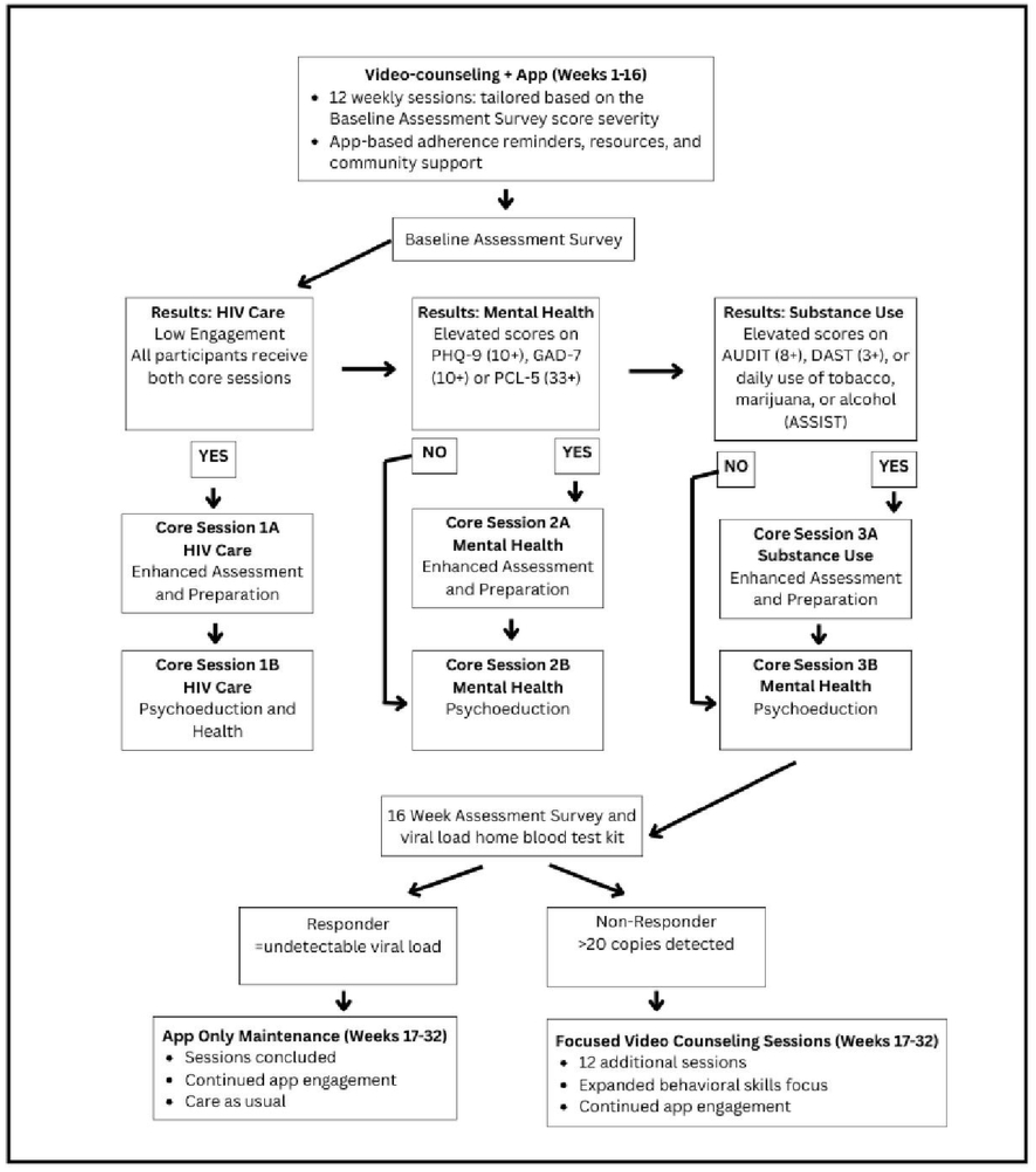
Study Design and Intervention Tailoring.

Overview of the method treatment strategy used in iVY, including baseline assessment, tailoring of intervention components based on mental health and substance use acuity, and assignment to Focused Sessions based on virologic response.

Given that all participants have experienced at least one detectable HIV viral load in the past 12 months (i.e., non-durable viral suppression), the intervention begins with two core HIV-focused sessions for each participant. Scores on measures of MH and SU described above guide the delivery of core sessions by tailoring the number of these sessions to individual needs. For example, participants with high MH symptom acuity on any baseline assessment receive two core MH sessions (Fig 1). Similarly, participants with high scores on any SU measure receive two SU sessions. The remaining 4-6 sessions (also known as menu sessions) are further tailored based on the individual’s needs and preferences (e.g., romantic and sexual relationships, subsistence needs, education, and vocation).

### WYZ App

Throughout the intervention, participants are encouraged to use WYZ to address various barriers to care (e.g., forgetting to take medications, social isolation, and the need for community resources). WYZ contains 3 main features: My Health, My Team, and My Community. Features in My Health include the ability to customize and track medication adherence over time, set medication reminders, and keep track of lab results. Features in My Team include access to location-specific community resources. Features in My Community include participation via an alias in a monitored forum with other YWH, access to a calendar of community events, and HIV-related news.

### Initial Session

After the enrollment visit, participants meet with their counselor individually for their initial session. As with all sessions in iVY, the initial session lasts 20-30 minutes. The session begins with an introduction and confirmation of the participant’s location and privacy. The objectives of this session are rapport building, intervention orientation, and discussion of the number of core sessions based on the participant’s baseline survey scores. The counselor clarifies roles and expectations (including clarifying that the iVY counselor should be seen as a health coach, not the participant’s therapist). The counselor and the participant also identify potential menu topics.

### Session Structure Overview

Following the initial session, the counselor delivers a series of core ‘A’ and ‘B’ sessions, as well as menu sessions to address individual needs. Consistent with the IMB model described above, each session type is based on one or two primary IMB components while allowing for overlap. Core ‘A’ sessions primarily target motivation, core ‘B’ sessions emphasize information and behavioral skills, and menu sessions focus on the behavioral skills.

### Core Sessions

Core sessions cover HIV care, MH, and SU, which are either motivation-focused (‘A’ sessions) or information-behavior-focused (‘B’ sessions). All participants receive both ‘A’ and ‘B’ HIV core sessions. Participants whose baseline survey results indicate higher acuity in MH or SU receive ‘A’ sessions related to MH or SU to enhance readiness to engage in care and behavior change. All participants receive the ‘B’ psychoeducation-focused session, which provides information and practical strategies on the topic.

All core sessions follow the same overall format (Fig 1). Core sessions begin with the counselor conducting a check-in, which includes confirming location and privacy, discussing one positive and one challenging experience from the previous week, and reviewing the past week’s goal (when applicable). The counselor then discusses the core session topic. For example, in the HIV core sessions, ‘A’ topic areas include current acceptance and understanding of HIV diagnosis, stigma-related beliefs, past experiences, and their impacts on current care, medication adherence, and lab work routines. The counselor reviews the baseline HIV knowledge assessment and provides feedback on the identified knowledge gaps. HIV related barriers are collaboratively identified. Using motivational interviewing, the counselor works with participants to enhance motivation to address these barriers in alignment with participants’ values and goals. Relevant resources are emailed or texted after the session. In the HIV core ‘B’ sessions, the counselor assesses the participant’s understanding of HIV care and knowledge gaps. Psychoeducation is provided on topics such as finding a supportive clinic, attending visits, HIV pharmacology, integrating medications into lifestyle, typical side effects, obtaining labs, and medical literacy to support sustained engagement in care. Additional useful resources are sent following the session. Similar to the check-in at the beginning of all core sessions, there is a check-out for each session, which elicits client feedback and provides a positive reflection on a strength and/or the client’s willingness to participate in the conversation.

### Menu Sessions

After completing the core sessions, the counselor and participant proceed through menu sessions, where a topic is selected collaboratively from a predetermined list of challenges that can influence an individual’s ART adherence. Menu sessions begin with a similar introduction to Core sessions (e.g., confirming the participant’s location and privacy, checking in on the previous week, reviewing progress toward their goal). Using a structured problem-solving approach, the counselor and the participant identify a barrier that impacts the participant’s overall health related to the menu topic, brainstorm potential solutions, evaluate them, collaboratively select the best option, develop a goal, and create a plan.

Menu sessions can be tailored to the individual’s needs and may include topics such as Current and Future Societal Concerns, Self-Identity and Disclosure, or Lifestyle Health. If the participant is experiencing a crisis (e.g., an acute challenge that does not clearly fit within the prescribed menu sessions), a “wildcard” session may be used. Counselors may also conduct a wildcard session after providing a safety assessment and simultaneous crisis intervention in the event of an MH crisis or a situation warranting mandated reporting.

### Final Session

The final session of the first 12-session counseling series emphasizes integration and reflection, helping participants consolidate learned skills, recognize progress, and strengthen motivation to continue engaging in care. In this session, the counselor reviews key material from previous sessions, focusing on the aspects the client found most beneficial while highlighting strengths and progress made throughout the intervention. Participants whose 16-week HIV viral load is detectable are offered the “Focused Sessions,” and if their viral load is undetectable, they continue using WYZ only (Fig 1).

### Focused Session Overview

The second set of 12 sessions, referred to as the Focused sessions, follows a similar format to the first 12, with an increased emphasis on addressing barriers to viral suppression through applied behavioral skills. Focused sessions center on the motivation and behavior components of the IMB model by using motivational interviewing and practical behavioral skills. The Focused session skills were selected to provide brief, practical behavioral strategies that could be delivered within a 20–30-minute session and directly applied to barriers affecting HIV care engagement. Skills were chosen based on their alignment with the behavioral skills component of the IMB model, relevance to the needs of youth living with HIV, and adaptability to diverse experiences. See Table 2 for a description of the Focused session topics and how motivation and behavioral skills are addressed for each.

Sessions have accompanying interactive worksheets informed by evidence-based therapeutic approaches such as Cognitive Behavioral Therapy, Dialectical Behavior Therapy, Acceptance and Commitment Therapy, and Motivational Interviewing [33–38]. Handouts are organized by skill topic and are emailed after the session as a resource to facilitate independent skill application. Example worksheets used during the Focused Sessions are available through the iVY intervention website (https://ivy.ucsf.edu/content/overview-of-intervention-delivery), which includes intervention materials and behavioral skills worksheets used throughout the intervention. As in the first 12 intervention sessions, Focused sessions are 20–30 minutes in length and begin with an initial session, followed by core sessions, a selection of menu sessions, and a final session. A wildcard session is also available for any immediate issues outside the core and menu session topics.

### Initial Focused Session

The initial Focused session begins with an overview of the similarities and differences from the first 12 intervention sessions. The counselor discusses the behavioral skills emphasis of these sessions, reviews the bio-psycho-social assessment from the 16-week survey, and elicits feedback about the participant’s experience with the first 16 weeks of intervention. To conclude, the counselor and participant identify and rank priority areas where the participant faces challenges in healthcare engagement to focus on in upcoming sessions.

### Core Focused Sessions

Following the initial Focused session, participants receive five core sessions delivered in sequential order: (1) Decisional Balance, (2) SMART Goals, (3) Action Plan, (4) Evaluate Outcomes, and (5) 7 Steps of Problem Solving. These sessions center on deepening the work done in the first 12 sessions by reinforcing and applying previously introduced behavioral skills. For example, in the first 16 weeks of the intervention, participants identify tangible goals. In the core Focused sessions, participants continue to work on goal-setting, using SMART (specific, measurable, attainable, relevant, timely) goals. After conducting the same check-in used in the first 16 weeks of the intervention, the counselor reviews the behavioral skill using an interactive, fillable worksheet (which includes basic information on the skill concept and a concrete example) via screenshare on the secure videoconferencing platform. Using the worksheet as a guide, the counselor reviews the content with the participant, allowing them to direct the process based on their understanding of the material. Psychoeducation on the skill is provided throughout, with the counselor offering clarification and addressing knowledge gaps as needed. The session proceeds by applying the skill to a collaboratively selected barrier that impacts treatment adherence or overall well-being. Participants’ desire to use the skill on their own is measured using importance and confidence rulers commonly used in Motivational Interviewing, which help the participant assess the perceived value of making a change and their self-efficacy to do so. The session concludes with strengthening motivation to use the behavioral skill in daily life.

The first core Focused session, which covers the decisional balance skill, serves as a model for the structure of all core sessions. The counselor begins by screensharing the worksheet (Fig 2) and discussing with the participant what is remembered about this concept from past sessions, identifying and addressing lingering areas of uncertainty. The counselor reviews the main points and a concrete example, such as exploring the benefits and costs of exercising three times a week. The counselor normalizes ambivalence about change and discusses how evaluating the benefits and costs before making a decision can be helpful. A relevant healthcare barrier is identified for applying the decisional balance skill to the participant’s own life (e.g., switching to injectable ART, attending a therapy group, or cutting down vaping). Using screenshare, the participant and counselor complete the decisional balance worksheet for the selected barrier. The counselor encourages the participant to choose the best option to focus on over the next week, identifying internal and external resources, skills, strengths, and past successes to support them in achieving the goal and practicing the skill. The current stage of change is assessed using the Importance and Confidence Rulers. The session ends with the participant setting a goal related to the skill covered.

**Fig 2.**
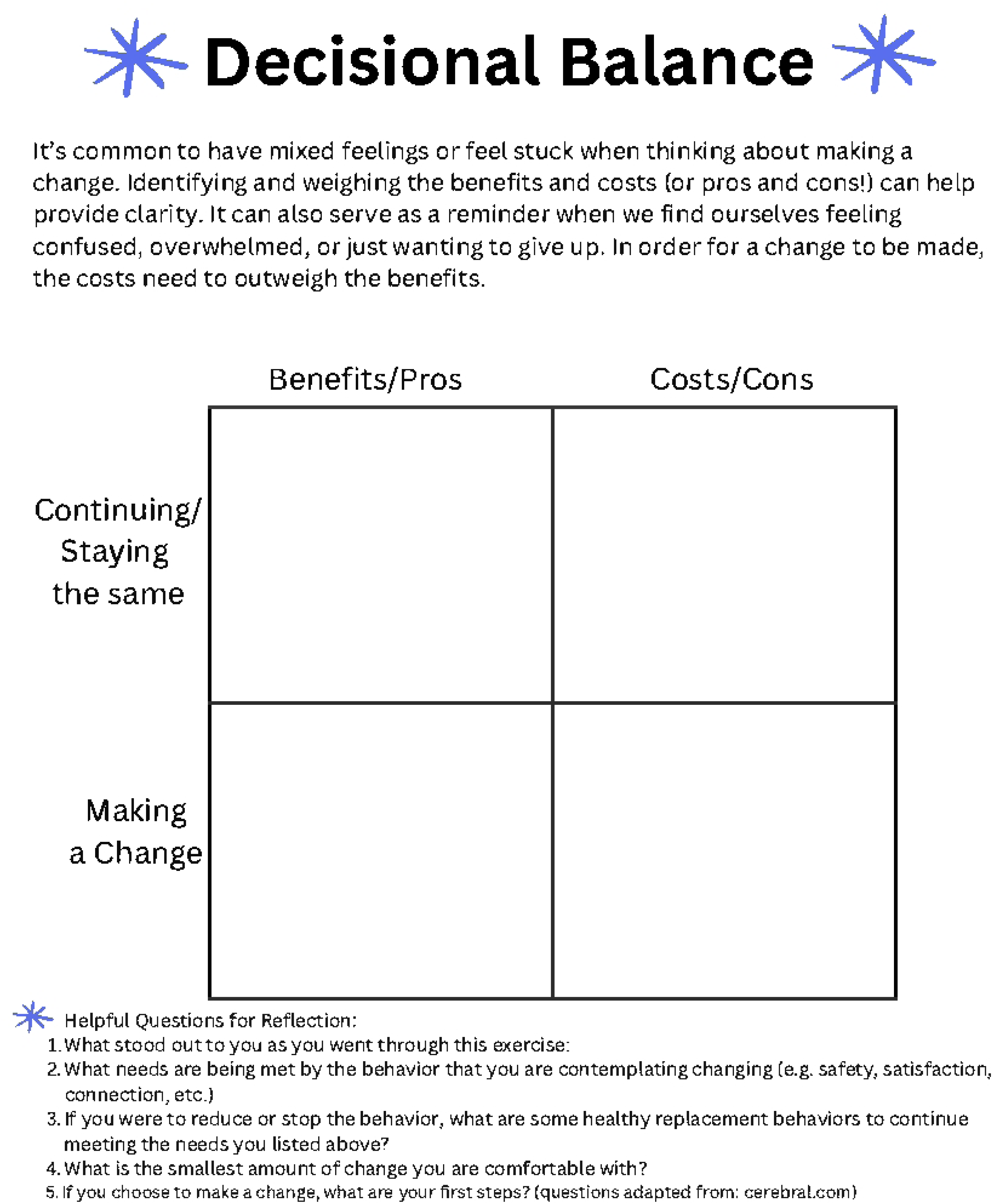
Decisional Balance Worksheet.

Example worksheet used during the Focused Sessions to evaluate the perceived advantages and disadvantages of behavior change and support decision-making related to HIV care engagement and overall health.

### Menu Focused Sessions

After the Core Focused sessions, the intervention continues with five menu Focused sessions that can be completed in any sequence. The counselor selects a session based on the participant’s identified focus area and barrier for that week. Menu-focused sessions follow the same format as core-focused sessions, with the key difference that the counselor introduces a new skill topic that complements but is not explicitly covered in the first 12 sessions. There are nine menu sessions to choose from: eight skills-focused sessions—Pleasurable Activities, Stress and Coping, Effective Communication, Boundaries, Practicing Acceptance, CBT (Thought, Emotion, Behavior) Connection, Identifying Values, Recognizing and Responding to Triggers, and the Motivation Enhancement session (see Table 2 for topic descriptions).

### Final Focused Session

The final Focused session highlights learnings throughout the intervention and creates a plan for ongoing goals. Previous sessions are reviewed, with the participant sharing the aspects that stood out to them. The counselor identifies and reinforces changes made during the intervention, noting successes and challenges, providing encouragement for those changes, recapping information on change strategies, and enhancing motivation for those changes. The counselor and participant identify ongoing goals and unmet needs that would benefit from continued care. Additional resources are provided to help the client connect to services or treatments needed for continued support.

## Discussion

iVY is a novel intervention for supporting YWH by providing a comprehensive approach to addressing the psychosocial and structural factors that may underpin the disparate rates of HIV incidence, decreased retention, and disparate rates of viral suppression that YWH experience compared to other age groups [4,5]. The intervention seeks to accomplish this by including MH, SU, and other youth-specific topics tailored to participants’ current barriers to healthcare engagement. Below, we present several guiding principles for delivering the intervention.

### Guiding Principles

The guiding principles of this video-based counseling intervention refer to the foundational values and clinical orientations that shape counselor engagement, ethical boundaries, and decision-making throughout the intervention. These principles emphasize clarity of role, ethical scope of practice, person-centered engagement, cultural responsiveness, and readiness-based support. Together, they guide counselors in fostering rapport, supporting autonomy, and promoting engagement without exceeding the intended counseling framework.

For many participants, this intervention serves as their first experience with counseling, which can lead to confusion with psychotherapy. During the initial session, it is essential to clarify that counseling differs from psychotherapy. While counselors recognize the personal nature of the discussions, they avoid delving too deeply into sensitive topics. If participants express readiness, counselors recommend additional MH resources to help participants find appropriate psychotherapy services.

Some participants exhibit impaired judgment or limited insight due to MH or SU challenges, or internalized stigma, making them less ready to acknowledge behavioral or life challenges. In these cases, Motivational Interviewing [24] and a strengths-based approach [39] provide valuable strategies to foster engagement and self-reflection, aligning with person-centered care rather than prematurely emphasizing problem-solving.

Culture should also be consistently considered when delivering iVY. According to the Centers for Disease Control and Prevention, Black/African Americans and Hispanic/Latinx people made up more than 70% of estimated new HIV infections in 2022 [3]. Therefore, iVY counselors incorporate cultural considerations into the ethical delivery of the intervention.

Recognizing the cultural diversity within these populations, the intervention acknowledges its roots in Western counseling ideologies and practices [40] while emphasizing the importance of culturally responsive counseling. To avoid homogenizing racial or ethnic groups, counselors adopt a trauma-informed person-centered approach [41] tailoring to each client’s unique experiences, cultural background, and values. This approach ensures the intervention is adaptable and respectful of participants’ identities and lived experiences. Counselors prioritize rapport-building while remaining sensitive to the cultural and systemic factors that may influence participants’ MH, SU, and engagement with healthcare services. By centering the individual, the intervention strives to create an inclusive and supportive environment for all participants.

### Strengths and Limitations

iVY has several key strengths that position it as a holistic and impactful intervention for people with HIV, specifically YWH. The distinctive approach uses a fully remote, technology-based format to effectively engage hard-to-reach YWH in a manner that aligns with their cultural norms regarding technology use. By meeting participants in their environment and positioning them as experts in their own needs, iVY empowers them to decide session topics and focus areas. The intervention’s broad range of topics is another strength, addressing the multiple factors influencing viral suppression and offering support for each. In addition, intervention content is adaptable, effectively identifying and addressing individual barriers to wellness engagement. Sessions are shorter (20–30 minutes) than those in traditional behavioral health interventions, which may also support sustainability. Additionally, the flexibility in scheduling the intervention has been crucial to participants’ engagement. To further support engagement, the counselor administering the intervention also offers regular drop-in hours, allowing participants to connect without advance scheduling. Practicing with an understanding of the potential need to adjust session times due to factors often outside of the participant’s control, such as unstable housing or variable work hours, allows participants flexibility to reschedule without breaking rapport. The availability of drop-in hours reflected an understanding of participants’ lived realities and served as a practical, low-barrier way to stay connected with youth whose schedules and circumstances were often unpredictable.

iVY has several limitations. Engaging YWH is an ongoing challenge, as the study targets not durably virally suppressed YWH. The study team endeavors to mitigate these challenges by offering flexible appointment times, allowing multiple reschedules, and providing bi-directional text messaging. Although the intervention remains responsive, some participants struggle to stay engaged, citing complex life circumstances, limited time and space, SU, and other barriers. While the use of technology as the primary means of communication aligns with YWH’s cultural norms and offers flexibility in intervention delivery, it also poses challenges. These include the need for access to a device and finding a private space for sessions. In addition, balancing engagement with the intervention when the topic is predetermined can be challenging at times. Wildcard sessions are applied in these circumstances to address issues requiring immediate attention.

## Conclusion

iVY integrates multiple levels of support—HIV care, MH, SU, and other psychosocial factors—into a cohesive, technology-based intervention tailored to the needs of YWH. By combining brief, structured video-based counseling with a mobile health platform and an adaptive treatment strategy, iVY addresses key barriers to care engagement in a flexible, accessible format. As outcomes from the ongoing RCT become available, this work provides a detailed and replicable description of a telehealth-based behavioral intervention and demonstrates a scalable model for integrating behavioral health and HIV care for YWH.

## Data Availability

This manuscript describes the development and structure of an intervention. No datasets were generated or analyzed for this study.

https://ivy.ucsf.edu/content/overview-of-intervention-delivery

## Acknowledgments

We would like to acknowledge feedback and collaboration from participants from previous studies, without which we could not have developed this intervention. Research coordinators Kristin Ming, Louis Smith, and Erin Moore provided invaluable support for this intervention.

## References

1. CDC. Estimated HIV incidence and prevalence. In: HIV Data [Internet]. 7 Feb 2025 [cited 9 Sept 2025]. Available: https://www.cdc.gov/hiv-data/nhss/estimated-hiv-incidence-and-prevalence.html

2. The White House. National HIV/AIDS Strategy for the United States 2022–2025. 2021.

3. CDC. Fast Facts: HIV in the US by Race and Ethnicity. In: HIV [Internet]. 27 June 2024 [cited 17 Dec 2025]. Available: https://www.cdc.gov/hiv/data-research/facts-stats/race-ethnicity.html

4. Allan-Blitz LT, Mena LA, Mayer KH. The ongoing HIV epidemic in American youth: challenges and opportunities. Mhealth. 2021;7: 1–13. doi:10.21037/mhealth-20-42

5. Kapogiannis BG, Koenig LJ, Xu J, Mayer KH, Loeb J, Greenberg L, et al. The HIV Continuum of Care for Adolescents and Young Adults Attending 13 Urban US HIV Care Centers of the NICHD-ATN-CDC-HRSA SMILE Collaborative. JAIDS Journal of Acquired Immune Deficiency Syndromes. 2020;84: 92–100. doi:10.1097/qai.0000000000002308

6. Kim SH, Gerver SM, Fidler S, Ward H. Adherence to antiretroviral therapy in adolescents living with HIV: systematic review and meta-analysis. AIDS. 2014;28: 1945–1956. doi:10.1097/QAD.0000000000000316

7. Zanoni BC, Mayer KH. The adolescent and young adult HIV cascade of care in the United States: exaggerated health disparities. AIDS Patient Care STDS. 2014;28: 128–135. doi:10.1089/apc.2013.0345

8. Casale M, Carlqvist A, Cluver L. Recent Interventions to Improve Retention in HIV Care and Adherence to Antiretroviral Treatment Among Adolescents and Youth: A Systematic Review. AIDS Patient Care and STDs. 2019;33: 237–252. doi:10.1089/apc.2018.0320

9. Dorfman M, Goldhammer H, Krebs D, Chavis NS, Psihopaidas D, Moore MP, et al. Interventions for Improving HIV Care Continuum Outcomes Among LGBTQ+ Youth in the United States: A Narrative Review. AIDS Patient Care and STDs. 2024;38: 358–369. doi:10.1089/apc.2024.0114

10. Laurenzi CA, du Toit S, Ameyan W, Melendez-Torres G, Kara T, Brand A, et al. Psychosocial interventions for improving engagement in care and health and behavioural outcomes for adolescents and young people living with HIV: a systematic review and meta-analysis. Journal of the International AIDS Society. 2021;24: e25741. doi:10.1002/jia2.25741

11. Mukerenge NF, Schmollgruber S, Klaas N. Health educational interventions for adolescents living with HIV: A scoping review. International Journal of Nursing Studies Advances. 2025;9: 100359. doi:10.1016/j.ijnsa.2025.100359

12. Gamarel KE, Brown L, Kahler CW, Fernandez MI, Bruce D, Nichols S. Prevalence and correlates of substance use among youth living with HIV in clinical settings. Drug and Alcohol Dependence. 2016;169: 11–18. doi:10.1016/j.drugalcdep.2016.10.002

13. Gonzalez JS, Batchelder AW, Psaros C, Safren SA. Depression and HIV/AIDS Treatment Nonadherence: A Review and Meta-analysis. JAIDS Journal of Acquired Immune Deficiency Syndromes. 2011;58: 181–187. doi:10.1097/QAI.0B013E31822D490A

14. Krumme AA, Kaigamba F, Binagwaho A, Murray MB, Rich ML, Franke MF. Depression, adherence and attrition from care in HIV-infected adults receiving antiretroviral therapy. Journal of Epidemiology and Community Health. 2015;69: 284–289. doi:10.1136/jech-2014-204494

15. Giovenco D, Muessig KE, Horvitz C, Biello KB, Liu AY, Horvath KJ, et al. Adapting technology-based HIV prevention and care interventions for youth: lessons learned across five U.S. Adolescent Trials Network studies. Mhealth. 2021;7: 21. doi:10.21037/mhealth-20-43

16. Saberi P, Stoner MCD, McCuistian CL, Balaban C, Ming K, Wagner D, et al. iVY: protocol for a randomised clinical trial to test the effect of a technology-based intervention to improve virological suppression among young adults with HIV in the USA. BMJ Open. 2023;13: e077676. doi:10.1136/bmjopen-2023-077676

17. Saberi P, McCuistian C, Agnew E, Wootton AR, Legnitto Packard DA, Dawson-Rose C, et al. Video-Counseling Intervention to Address HIV Care Engagement, Mental Health, and Substance Use Challenges: A Pilot Randomized Clinical Trial for Youth and Young Adults Living with HIV. Telemedicine Reports. 2021;2: 14–25. doi:10.1089/tmr.2020.0014

18. McCuistian C, Wootton AR, Legnitto-Packard D, Gruber VA, Dawson-Rose C, Johnson MO, et al. Addressing HIV care, mental health and substance use among youth and young adults in the Bay Area: Description of an intervention to improve information, motivation and behavioural skills. BMJ Open. 2021;11: 1–9. doi:10.1136/bmjopen-2020-042713

19. Erguera XA, Johnson MO, Neilands TB, Ruel T, Berrean B, Thomas S, et al. WYZ: a pilot study protocol for designing and developing a mobile health application for engagement in HIV care and medication adherence in youth and young adults living with HIV. BMJ Open. 2019;9: e030473. doi:10.1136/bmjopen-2019-030473

20. Saberi P, Lisha NE, Erguera XA, Hudes ES, Johnson MO, Ruel T, et al. A Mobile Health App (WYZ) for Engagement in Care and Antiretroviral Therapy Adherence Among Youth and Young Adults Living With HIV: Single-Arm Pilot Intervention Study. JMIR Form Res. 2021;5: e26861. doi:10.2196/26861

21. Fisher JD, Fisher WA. Changing AIDS-Risk Behavior. 1992;I: 455–474.

22. Fisher JD, Amico KR, Fisher WA, Harman JJ. The Information-Motivation-Behavioral Skills model of antiretroviral adherence and its applications. Curr HIV/AIDS Rep. 2008;5: 193–203. doi:10.1007/s11904-008-0028-y

23. Lukens EP, McFarlane WR. Psychoeducation as Evidence-Based Practice: Considerations for Practice, Research, and Policy. Brief Treatment and Crisis Intervention. 2004;4: 205–225. doi:10.1093/brief-treatment/mhh019

24. Miller WR, Rollnick S. The Spirit of Motivational Interviewing. Motivational Interviewing : Preparing People for Change. 2013; 14–24.

25. Nezu, AM, Nezu CM, D’Zurilla T. Problem-solving therapy: A treatment manual. Co. SP, editor. New York, NY; 2013.

26. Chenneville T, Gabbidon K, Drake H, Rodriguez C. Comparison of the utility of the PHQ and CES-D for depression screening among youth with HIV in an integrated care setting. Journal of Affective Disorders. 2019;250: 140–144. doi:10.1016/j.jad.2019.03.023

27. Spitzer RL, Kroenke K, Williams JBW, and the Patient Health Questionnaire Primary Care Study Group. Validation and Utility of a Self-report Version of PRIME-MDThe PHQ Primary Care Study. JAMA. 1999;282: 1737–1744. doi:10.1001/jama.282.18.1737

28. Spitzer RL, Kroenke K, Williams JBW, Löwe B. A brief measure for assessing generalized anxiety disorder: The GAD-7. Archives of Internal Medicine. 2006;166: 1092–1097. doi:10.1001/archinte.166.10.1092

29. Lang AJ, Wilkins K, Roy-Byrne PP, Golinelli D, Chavira D, Sherbourne C, et al. Abbreviated PTSD Checklist (PCL) as a guide to clinical response. General Hospital Psychiatry. 2012;34: 332–338. doi:10.1016/j.genhosppsych.2012.02.003

30. Johnson JA, Lee A, Vinson D, Seale JP. Use of AUDIT-Based Measures to Identify Unhealthy Alcohol Use and Alcohol Dependence in Primary Care: A Validation Study. Alcoholism: Clinical and Experimental Research. 2013;37: 253–259. doi:10.1111/j.1530-0277.2012.01898.x

31. Skinner HA. The drug abuse screening test. Addictive Behaviors. 1982;7: 363–371. doi:10.1016/0306-4603(82)90005-3

32. Balfour L, Kowal J, Tasca GA, Cooper CL, Angel JB, MacPherson PA, et al. Development and psychometric validation of the HIV Treatment Knowledge Scale. AIDS Care - Psychological and Socio-Medical Aspects of AIDS/HIV. 2007;19: 1141–1148. doi:10.1080/09540120701352241

33. Beck JS. Cognitive Behavior Therapy, Second Edition: Basics and Beyond. New York, NY: The Guilford Press; 2011.

34. CBT Triangle Worksheet. In: Therapist Aid [Internet]. [cited 19 Dec 2025]. Available: https://www.therapistaid.com/therapy-worksheet/cbt-triangle

35. Decisional Balance Worksheet. In: Motivational Interviewing Network of Trainers (MINT) [Internet]. 2011 [cited 19 Dec 2025]. Available: https://motivationalinterviewing.org/decisional-balance-worksheet

36. Gordon T, Borushok J, Polk K. The ACT Approach. PESI Publishing Media; 2017. Available: https://www.pesi.com/item/the-act-approach-42236

37. Miller AL, Rathus JH, Dexter-Mazza ET, Brice CS, Graling K. DBT for Adolescents. 2nd edition. Dialectical Behavior Therapy in Clinical Practice: Second Edition: Applications across Disorders and Settings. 2nd edition. New York, NY: The Guilford Press; 2021. pp. 345–365. Available: https://www.guilford.com/books/Dialectical-Behavior-Therapy-in-Clinical-Practice/Dimeff-Rizvi-Koerner/9781462552641?srsltid=AfmBOoqL1lmAqAL1pzCtAPf3npUHz4NF8qKBvyGwP8A0kI00V3zatdKW

38. Snyder-Sclater D. Enhancing Team Based Care in the Outpatient Setting: Problem Solving Techniques. 2021. Available: https://micmt-cares.org/sites/default/files/2021-12/PST%20training%2012-15-21%20final.pdf

39. Pulla V. Strengths-Based Approach in Social Work: A distinct ethical advantage. International Journal of Innovation. 2017;3.

40. Cheung FM. Deconstructing Counseling in a Cultural Context. The Counseling Psychologist. 2000;28: 123–132. doi:10.1177/0011000000281008

41. Yadav G, McNamara S, Gunturu S. Trauma-Informed Therapy. StatPearls [Internet]. StatPearls Publishing; 2024. Available: https://www.ncbi.nlm.nih.gov/books/NBK604200/

